# Influence of organisational culture on knowledge generation and application within Learning Health Systems: a scoping review protocol (Organisational culture and Learning Health Systems)

**DOI:** 10.1101/2025.05.14.25327638

**Authors:** Seo Yeon Yoon, Georgia Bercades, Matthew G. Wilson, Yogini H Jani

## Abstract

**Background:** A learning health system (LHS) is a framework within healthcare that continuously enables improvement by incorporating knowledge generation into practice, leveraging routine data to optimise patient outcomes. Organisational culture, encompassing workplace values and traditions, is crucial for LHSs, allowing for integration of various elements within the system and ensuring cohesive functionality. Without a supportive organisational culture, a LHS may face challenges in achieving positive results, even with well-functioning individual components. Despite its recognised importance in LHS literature, the impact of organisational culture on LHS success remains unclear. This review aims to bridge this research gap.

**Objectives:** The scoping review will address the question: How does organisational culture influence the generation and application of knowledge within a LHS?

**Methods:** This scoping review will follow Joanna Briggs Institute scoping review guidelines. Studies examining organisational culture within LHSs will be included, whilst those examining organisational culture outside a LHS framework not. Screening will be conducted using Rayyan, with title and abstract screening, followed by full-text review. A calibration exercise will be undertaken to ascertain agreement of eligibility criteria, after which the second reviewer will independently screen 10% of the studies during the title and abstract screening. A third reviewer will resolve any disagreements. Data extraction will use a standardised data information sheet, and the selection process and results of chosen studies will be recorded using the Preferred Reporting Items for Systematic reviews and Meta-Analyses extension for Scoping Reviews (PRISMA-ScR) flow diagram. Findings will be presented through a narrative summary using thematic analysis.

**Ethics and Dissemination:** This review does not require an ethics approval. This protocol is registered to Open Science Framework and is publicly available at https://doi.org/10.17605/OSF.IO/K4EC3.

## Introduction

### Background and Rationale

**What is a learning health system?**

A learning health system (LHS) can be defined as a framework in which patient outcomes and experiences are continually improved by applying sciences, informatics, incentives, and cultural frameworks to generate and utilise knowledge in the delivery of care(1). The concept of a LHS was introduced by the Institute of Medicine (IoM; now the National Academy of Medicine) in 2007 through a workshop to identify issues most important in bridging the gap between research and clinical decisions. It also aimed to create a system where knowledge generation and application are embedded into each stage of healthcare delivery(2). Examples of this framework include the LHS within University of Montreal using integrated data to build clinical decision-making tools(3), and the University of Pittsburgh Medical Centre’s rapid learning health system, constructed in response to the Covid-19 pandemic(4).

Based on existing research of LHSs, this review proposes three defining elements of LHS (Figure 1) that need to work symbiotically. These will be used to determine whether an organisation successfully follows the LHS framework. These three elements: 1) continuous knowledge generation and application aimed at 2) improving patient outcomes, and 3) the sustained integration of 1) and 2) at every stage of care to emphasise a culture aligned with continuous learning and improvement, can be utilised to assess the LHS status of organisations (Table 1).

**Figure 1:**
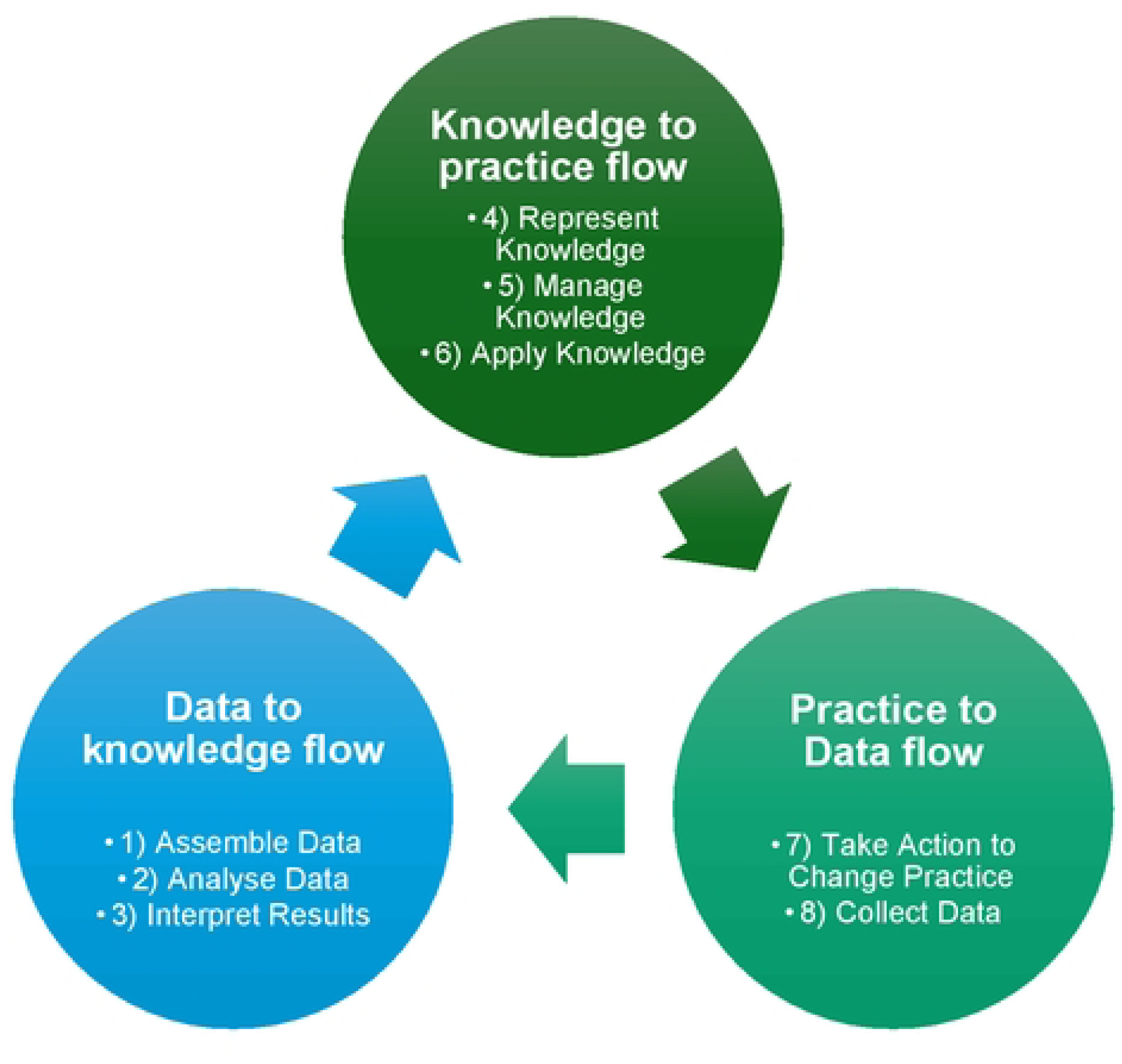
Learning health cycle of a LHS adapted from (1).

**Table 1:**
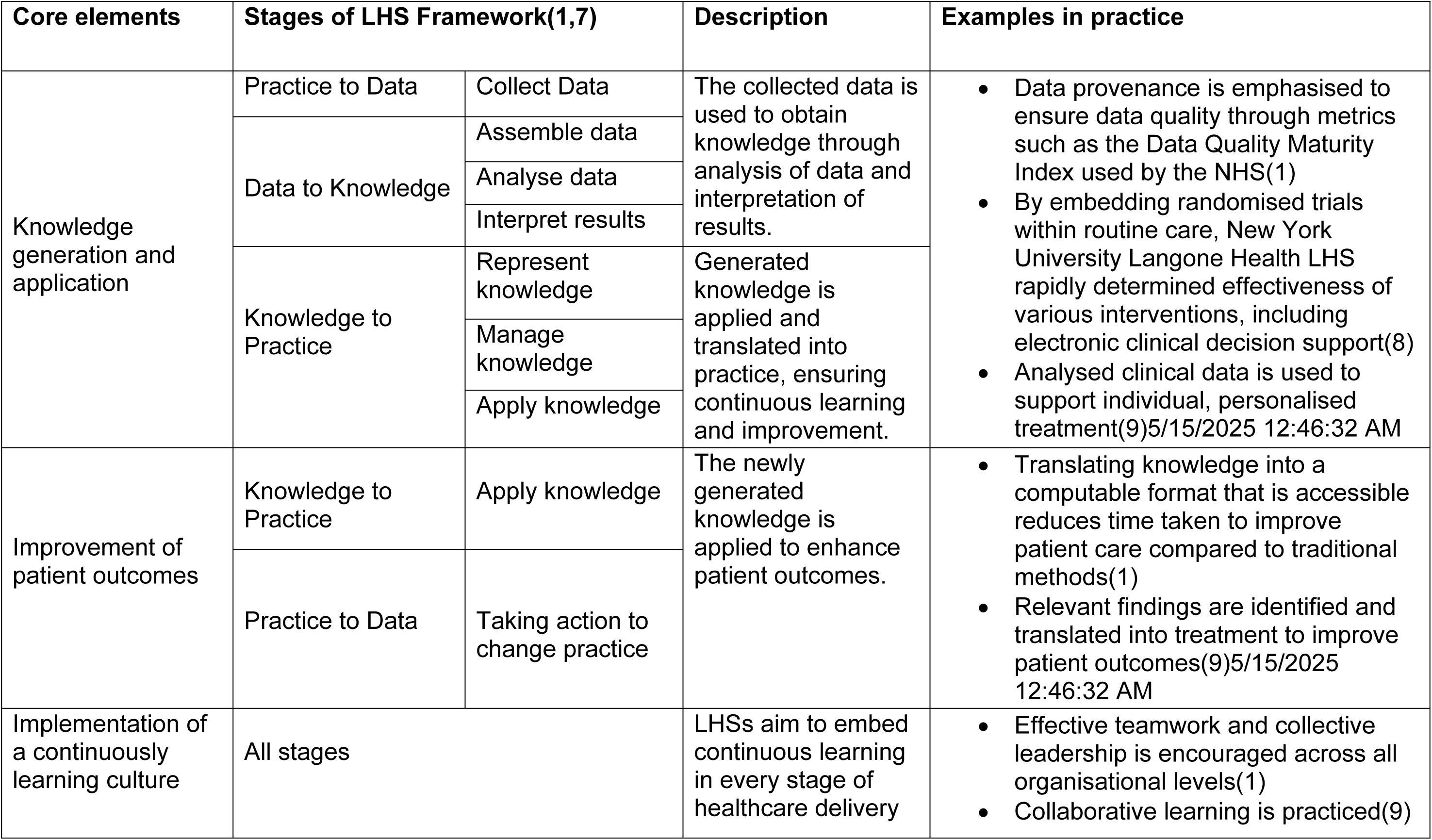
Core elements within LHSs.

LHSs rely on data and knowledge generated within healthcare to improve patient experiences and create adaptative healthcare. However, the application of the data generated by health information technologies alone may not be sufficient in improving patient care as organisational culture remains an important factor(5).

In 2018, the UK National Health Service (NHS) launched a framework, IMPACT-Improving Patient Care Together, to promote continuous improvement(6). One of the five components of IMPACT, “investing in people and culture”, highlights that healthy cultures in NHS organisations are essential to delivering high-quality patient care(6).

Reflected in the values, norms, and beliefs of the individuals that are part of an organisation (e.g. clinicians working in a hospital), organisational culture is an important part in understanding the dynamics, inner workings and effectiveness of a healthcare network, such as LHSs(10,11).

Although culture is presented as one of the key dimensions of LHSs, findings from existing research underscore a lack of understanding of the role of organisational culture in facilitating the technological implementations that serve to support LHSs(5,12). Moreover, current LHS literature highlights despite advantages of examining organisations as complex interdependent systems to LHS development, involvement of organisational culture, such as the integration of socioecological engagement and the involvement of stakeholders(13), are generally poor. The Learning Healthcare Project reiterates this point by stating although culture is clearly an important factor, there is difficulty in determining the relationship between culture and LHSs due to the multidisciplinary nature and complexity of the system(16). Current literature highlights the need for further exploration on the relationship, but despite these evidence gaps, the extent and breadth of the existing literature pertaining to the role of organisational culture within LHS remains unknown. This scoping review aims to bridge this literature gap for future research with the guidance of the review questions.

### Defining organisational culture

Existing research on organisational culture in relation to patient safety demonstrates inconsistency in definition and usage of the term. In this scoping review, the following definition of organisational culture will be used:

> “Organizational culture is the pattern of basic assumptions which a given group has invented, discovered, or developed in learning to cope with its problems of external adaptation and internal integration, which have worked well enough to be considered valid, and therefore, to be taught to new members as the correct way to perceive, think, and feel in relation to those problems.”

> (*Organizational Culture: A Dynamic Model*, Schein, 1983)

As one of the most popular definitions of organisational culture(18), Schein’s work aims to provide clarity to the concept of culture by using a model(19). This model is used as a framework to explore organisational culture across three levels: 1) artefacts and behaviours, 2) espoused values, and 3) basic underlying assumptions(17–19)(Figure 2). This model allows for a structured approach to understanding organisational culture and emphasises the importance of cultural awareness in creating a successful working environment(20). It offers an in-depth understanding of how culture operates theoretically and practically, which aligns with the ultimate goal of continuous learning within a LHS. Therefore, this definition accommodates this review’s aims in understanding the influence of organisational culture within a LHS.

**Figure 2:**
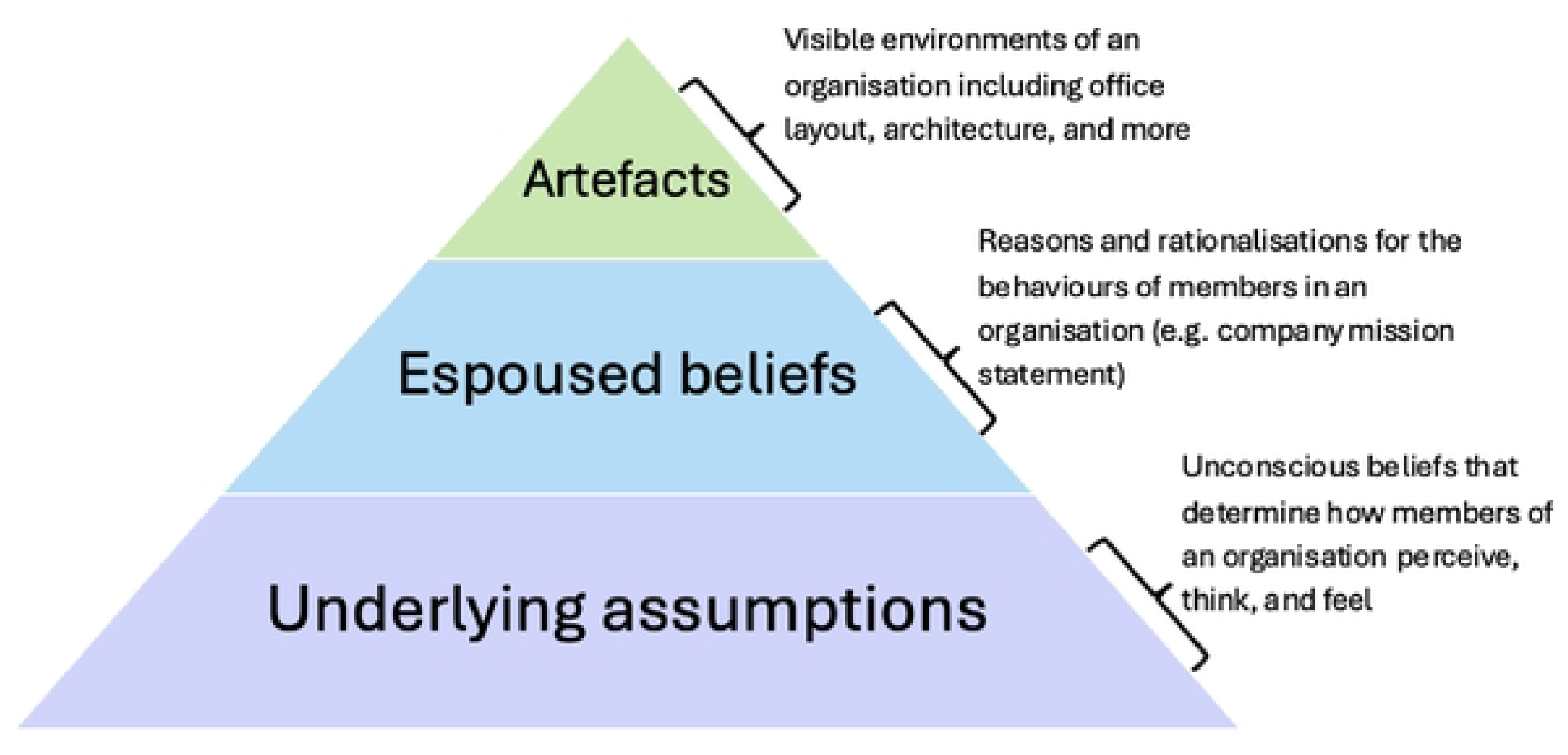
Schein’s model of organisational culture adapted from Doan(21)(22).

Table 2 illustrates how each aspect of organisational culture will be evaluated within this study.

**Table 2:**
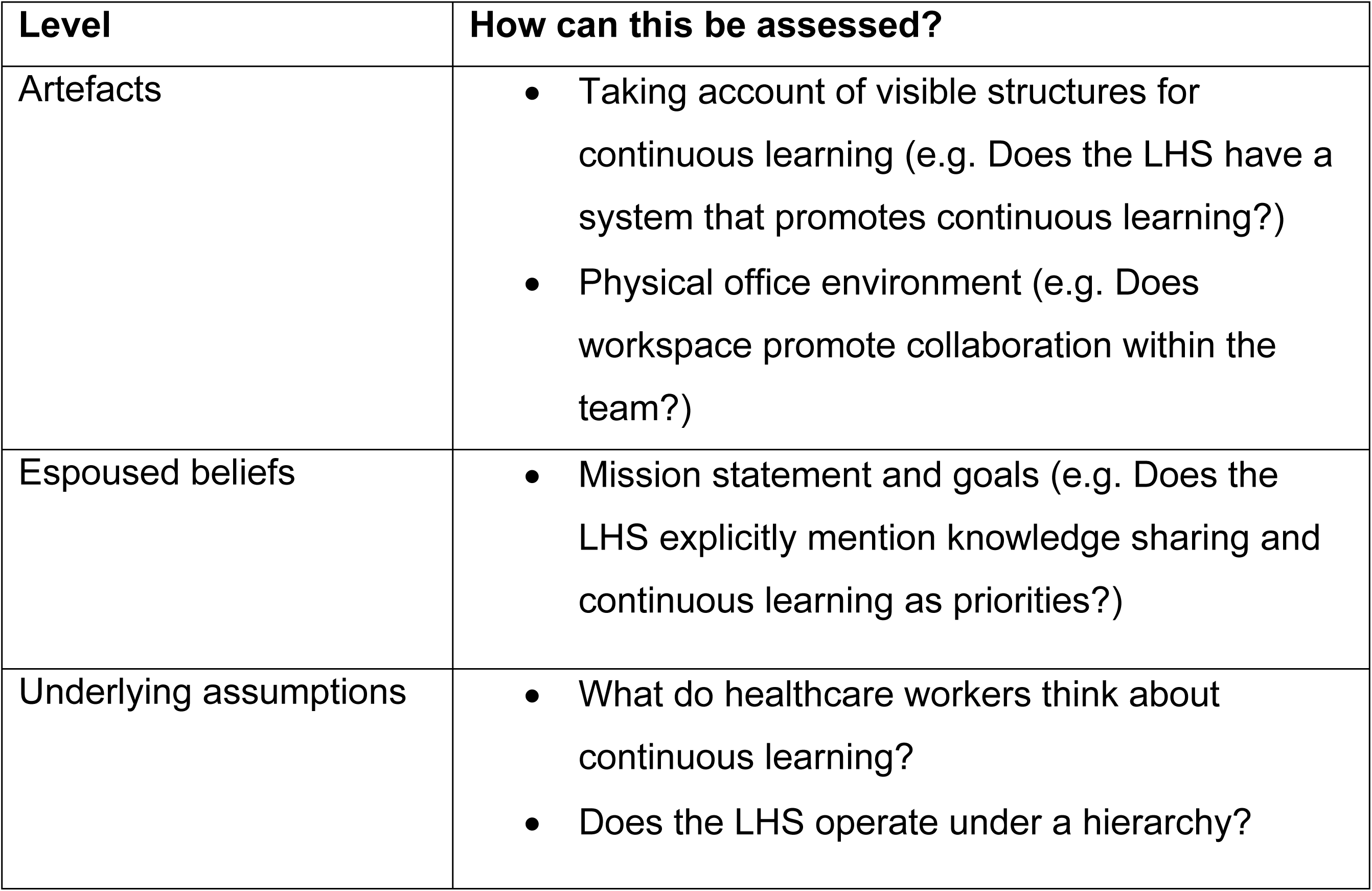
Applying Schein’s Organisational Culture model to LHSs.

| Level | How can this be assessed? |
| --- | --- |
| Artefacts | • Taking account of visible structures for continuous learning (e.g. Does the LHS have a system that promotes continuous learning?)<br>• Physical office environment (e.g. Does workspace promote collaboration within the team?) |
| Espoused beliefs | • Mission statement and goals (e.g. Does the LHS explicitly mention knowledge sharing and continuous learning as priorities?) |
| Underlying assumptions | • What do healthcare workers think about continuous learning?<br>• Does the LHS operate under a hierarchy? |

### Research Questions

The objective is to identify and examine how the current literature reports the influence of organisational culture on LHSs. The research question was developed using the population, concept, context (PCC) framework as recommended by Joanna Briggs Institute (JBI) (Table 3)(23). A preliminary search of PubMed in 2025 found no existing scoping reviews on this topic.

**Table 3:** The Population, Concept, Context (PCC) framework used to form the research question following the JBI framework for scoping review.

| **PCC Framework** |  |
| --- | --- |
| Population | User groups within LHS |
| Concept | Relationship between organisational culture and knowledge generation and application within LHSs |
| Context | LHS or system synonymous |

### Primary research question: How does organisational culture influence the generation and application of knowledge within a LHS?

The following sub questions further explore the research question:

- How is organisational culture defined within healthcare and LHS?
- What are the key cultural characteristics that enable and/or hinder knowledge generation within a LHS?
- What specific cultural attributes are associated with improving patient care within a LHS?
- How does the influence of organisational culture differ across LHSs?

## Methods

### Study Design

This scoping review will follow the JBI scoping review framework, and the Preferred Reporting Items for Systematic reviews and Meta-Analyses extension for Scoping Reviews (PRISMA-ScR) checklist(23,24).

### Eligibility criteria

#### Population

Users of LHSs, as primary characters contributing to organisational settings, such as clinicians, patients, administrators, stakeholders, and/or any other organisational units within a LHS will be the focus of the research population. The population will not be limited to specific subgroups within healthcare settings. Studies from all healthcare settings, such as clinics or hospitals, will be included to gain a holistic understanding of the relationship between knowledge generation and application within diverse LHSs.

#### Concept

The relationship between organisational culture and generation and application of knowledge will be the concept of this scoping review. The review will assess how organisational culture shapes knowledge generation and application, and if specific cultural elements impact patient care outcomes and knowledge utilisation.

#### Context

LHSs will serve as context for this research question. The review will include a wide range of organisations (e.g. hospitals, public health systems) that function as successful LHSs to best captivate a widespread context to answer the research question(s). As the review will include a large variety of different organisations, the type of LHSs (e.g. multinational organisations or local secondary care practice) will be specified during the data extraction process. Following Friedman(25)’s differentiation of LHSs from other cyclic improvement systems, LHS can be characterised as a continuously learning cycle that focuses on improving patient health outcomes through generation and application of knowledge, analysis of data, implementation of research findings, and engagement of stakeholders within a multi-collaborative organisational setting. This characterisation of LHS, along with the framework and core values discussed in Table 1, will be considered when reviewing and selecting studies that do not explicitly state an organisation to be a LHS. A sensitivity analysis will be conducted to ensure literature regarding LHS published before 2007 is not classified using different terminology.

### Search strategy

#### Search terms

The search strategy was developed using the two focused concepts of this review: LHS and organisational culture. To ensure the most thorough search, synonymous terms were explored using various methods. MeSH terms discovered during the initial search of existing literature were analysed, and the terms directly related to the two concepts were compiled. To ensure the concept of LHS is fully captured, synonyms outlined on the Learning Healthcare Project website were searched through Ovid (Medline and Embase).

The final search terms developed for this review are displayed in Table 4.

**Table 4:** finalised search terms for LHS and organisational culture.

| **Concept** | **Search Terms** |
| --- | --- |
| Learning Health System | learn* health* system* OR learning healthcare system* OR learning health systems OR learning health communit* OR system* learn* health* OR health* system* learn* OR data hub* OR living hub* OR innovation informatics hub* OR learning network* OR learning laborator* OR learning environment* OR community clinician participatory data healthcare research OR data driven improvement initiative* OR interventional informatics OR practice based data network* OR circular data driven healthcare OR rapid learning health system* |
| Organisational Culture | cultur* organi?ation* OR organi?ational decision making OR corporate cultur* OR cultur* corporate OR share* value* OR social value* OR social belief* OR safety cultur* OR cultur* OR organisational culture OR organizational culture OR professional culture* |

The search strategy will aim to find peer reviewed publications, as well as other reports, including grey literature. Multiple databases, including specific journals will be searched, as presented below:

- PubMed
- Medline
- Cochrane Library
- HMIC (Health Management)
- Embase (Ovid)
- Scopus
- Cumulative Index to Nursing and Allied Health Literature (CINAHL)
- PROSPERO
- Learning Health Systems Journal (Wiley)

Multiple large databases have been selected due to geographical distribution and types of research varying by database. The inclusion of grey literature is due to the estimation of limited number of resources available on the subject. The limited research on this topic may also lead to publication bias, which will be assessed by looking for positive and negative outcomes of the research question within literature. The grey literature will include health organisations or healthcare associations (e.g. Learning Healthcare Project website) that offer guidelines or frameworks related to LHSs, policy documents, government websites and white papers.

As the search terms constructed were based on the PubMed database, it is possible that adjustments to search terms may be required to permit searching across other databases. The search terms will be accordingly adjusted to each database, including but not limited to changes in spelling and truncation methods.

### Study Selection

Study selection will follow three stages: pilot calibration exercise, title and abstract screening, and full-text screening. As per the JBI scoping review guidelines(26), multiple (three) reviewers will be involved in the screening process to ensure consistency and reliability during the study selection process. To further assist the screening process, Rayyan, a web tool designed to help with knowledge synthesis(27), will be utilised. A pilot calibration exercise will be conducted, in which the two reviewers (SY, GB) independently screen 10% of the studies, to ensure 90% or above agreement of the inclusion and exclusion criteria (detailed in Table 5) (28). Inclusion criteria may need reviewing if reviewers do not come to agreement on 90% or more of the studies(28). Once this process is finalised, the second reviewer (GB) will independently screen 10% of the studies during the title/abstract screening and full-text review. In case of disagreement between reviewers, a third reviewer will make a final decision. The results of the study selection will be reported in the scoping review using the PRISMA-ScR flowchart(24). All selected literature will be stored and organised on Microsoft Excel.

**Table 5:** Inclusion and Exclusion Criteria.

|  | **Inclusion** | **Exclusion** |
| --- | --- | --- |
| Study Design | Any study designs, including those found through grey literature, case reports, descriptive reports, and reviews | N/A |
| Population | Any registered organisations and/or systems that follow the framework of LHSs | Organisations and/or systems that do not follow a LHS framework |
| Intervention | Literature aimed at understanding the influence of organisational culture, or a culture within organisational culture (i.e. professional culture) | Studies that do not focus on exploring the effects of organisational culture (e.g. learning health system and consent) |
| Outcome | Studies that outline the influence of culture in maintaining or building a LHS or system synonymous (e.g. using validated cultural instruments) | Studies that do not explore the influence of culture within their research |
| Time period | Studies from 2007 onwards (based on study start date) |  |

### Data Extraction

Data extraction will be performed using an extraction information sheet, using the PCC framework as a guide, to achieve the research objectives(29).

Given this review uses Schein’s definition of organisational culture, the model will also be used to guide the data extraction process. Artefacts, espoused beliefs, and basic assumptions of LHSs will be identified within the selected studies for those that do not use a validated cultural measurement tool. If a study has measured the influence of culture as part of their research using an alternative validated tool, the direct findings from the study will be extracted and mapped to Schein’s model.

The core elements stated in Table 1 will be referred to when assessing successful knowledge generation and application and subsequent improvements in patient care. Studies will be evaluated against key metrics aligned with the LHS concepts of patient-focused care and continuous learning. This may include quantitively (e.g. quantifying results from patient satisfaction survey) or qualitatively (e.g. exploring findings of patient interviews) measuring patient outcomes, examining disparities in patient experiences across different populations, investigating the number of new clinical guidelines and/or protocols produced, evidence of research outputs such as publications or new data, and use of feedback loops to improve future research for each LHS.

Data items for extraction will include the following:

- Author
- Title
- Country/region
- Aims
- Study design
- Population

- User groups within LHS
- Type of LHS (i.e. specify if system is multinational organisation or local secondary care LHS)
- Definition used for
- LHS (or a system synonymous)
- organisational culture
- Use of a validated cultural framework
- The Schein’s model

- Artefacts
- Espoused beliefs
- Underlying assumptions
- Key metrics of patient care and knowledge generation.

### Data Analysis and Presentation

The results of the selection process will be recorded according to the PRISMA-ScR flow diagram(24). The findings within the selected studies will be analysed to confirm alignment with the aims of this review and will be presented as a narrative summary.

The results of the data extraction will then be analysed thematically to examine specific cultural attributes that are associated with improving patient care within a LHS, and how the influence of organisational culture differs across LHSs. Thematic analysis, being a flexible method for identifying themes and patterns, provides an open opportunity to explore prevalence and themes in a number of ways through searching across a dataset(30). The review will follow Braun and Clarke’s six phases of analysis for conducting thematic analysis outlined in Table 6(30).

**Table 6:** Phases of thematic analysis(30).

| **Phase** | **Description of process** |
| --- | --- |
| 1. Familiarising yourself with data | Reading and re-reading data, noting down initial ideas |
| 2. Generating initial codes | Systematically coding noteworthy features of data across entire data set, collating data relevant to each code |
| 3. Searching for themes | Organising codes into potential themes, gathering all relevant data for each |
| 4. Reviewing themes | Examining themes in relation to each code and then entire data set to generate thematic “map” of the analysis |
| 5. Defining and naming themes | Analysis to refine each theme, the overall story the analysis tells, and generating clear names and definitions for each theme |
| 6. Producing the report | Selecting compelling extract examples, final analysis of selected extracts, relating back of the analysis to the research question to produce report of the analysis |

### Ethics and Dissemination

This review does not require an ethics approval. This protocol has been registered to Open Science Framework and is publicly available at https://doi.org/10.17605/OSF.IO/K4EC3.

### Status and timeline of study

Start date: March 2025 End date: August 2025

### Conclusion

This scoping review will highlight how organisational culture can affect LHS operation and success and identify gaps within current literature. The outcome of this review will identify and categorise characteristics of organisational culture that influence knowledge generation and application and improvements in patient care within a successful LHS. This review will provide evidence to support LHS development, directly contributing to improving the healthcare system.

## Author’s contributions

SY led the writing and conceptualisation of this review on exploring the effects of organisational culture on LHS development. MGW and YHJ contributed to supervision, the study concept, methodology and critical revisions of the manuscript. They will act as a third reviewer if needed. GB will contribute as a second reviewer. All authors reviewed the final manuscript prior to submission.

## Data Availability

No datasets were generated or analysed during the current study. All relevant data from this study will be made available upon study completion.

## Acknowledgements

SY would like to thank Laura Russell, a clinical support librarian at the Royal Free Hospital Medical Library at UCL for assisting in finalising the search strategy for this review.

## Supporting Information

- PRISMA-ScR checklist
- Search strategy for Embase and Medline

## Notes

**Funding:** This study is supported by the NIHR Central London Patient Safety Research Collaboration (CL-PSRC), reference number NIHR 204297. The views expressed are those of the author’s and not necessarily those of the NIHR or the Department of Health and Social Care. The authors Seo Yeon Yoon, Georgia Bercades and Dr Matthew Wilson are funded by the NIHR Central London Patient Safety Research Collaboration (CL PSRC). Dr Yogini H Jani is co-lead for Safer Evidence theme and Equality, Diversity & Inclusion lead for the NIHR CL PSRC, reference number NIHR204297. The views expressed are those of the author(s) and not necessarily those of the NIHR or the Department of Health and Social Care. This study forms parts of Seo Yeon Yoon’s PhD program.

### Competing Interest Statement

The authors have declared no competing interest.

### Funding Statement

Yes

### Author Declarations

This review does not require an ethics approval.

